# Chinese Public Attention to COVID-19 Epidemic: Based on Social Media

**DOI:** 10.1101/2020.03.18.20038026

**Authors:** Yuxin Zhao, Huilan Xu

## Abstract

**Background:** Since the new coronavirus epidemic in China in December 2019, information and discussions about COVID-19 have spread rapidly on the Internet and have quickly become the focus of worldwide attention, especially on social media.

**Objective:** This study aims to investigate and analyze the public’s attention to COVID-19-related events in China at the beginning of the COVID-19 epidemic in China (December 31, 2019, to February 20, 2020) through the Sina Microblog hot search list.

**Methods:** We collected topics related to the COVID-19 epidemic on the Sina Microblog hot search list from December 31, 2019, to February 20, 2020 and described the trend of public attention on COVID-19 epidemic-related topics. ROST CM6.0 (ROST Content Mining System Version 6.0) was used to analyze the collected text for word segmentation, word frequency, and sentiment analysis. We further described the hot topic keywords and sentiment trends of public attention. We used VOSviewer to implement a visual cluster analysis of hot keywords and build a social network of public opinion content.

**Results:** The study has four main findings. First, we analyzed the changing trend of the public’s attention to the COVID-19 epidemic, which can be divided into three stages. Second, the hot topic keywords of public attention at each stage are slightly different. In addition, the emotional tendency of the public toward the COVID-19 epidemic-related hot topics has changed from negative to neutral, with negative emotions weakening and positive emotions increasing as a whole. Finally, we divided the COVID-19 topics with the most public concern into five categories: new COVID-19 epidemics and their impact; (2) frontline reporting of the epidemic and prevention and control measures; (3) expert interpretation and discussion on the source of infection; (4) medical services on the frontline of the epidemic; and (5) focus on the global epidemic and the search for suspected cases.

**Conclusions:** This is the first study of public attention on the COVID-19 epidemic using a Chinese social media platform (i.e., Sina Microblog). Our study found that social media (e.g., Sina Microblog) can be used to measure public attention to public health emergencies. During the epidemic of the novel coronavirus, a large amount of information about the COVID-19 epidemic was disseminated on Sina Microblog and received widespread public attention. We have learned about the hotspots of public concern regarding the COVID-19 epidemic. These findings can help the government and health departments better communicate with the public on health and translate public health needs into practice to create targeted measures to prevent and control the spread of COVID-19.

## Introduction

Pneumonia caused by the novel coronavirus (COVID-19) is a new infectious disease that is mainly transmitted by respiratory droplets and contact and is generally infectious to human beings^1^. On January 11, 2020, after pathogenic nucleic acid testing (NAT), China reported 41 cases of pneumonia infected with the novel coronavirus (SARS-CoV-2) for the first time^2^, which was the world’s first reported human infection with the novel coronavirus. On January 30, 2020, the World Health Organization listed the novel coronavirus epidemic as a Public Health Emergency of International Concern (PHEIC) ^3^. As of February 20, 2020, a total of 75,465 confirmed cases and 2,236 deaths have been reported in mainland China ^4^. The novel coronavirus has caused great challenges and threats to public health in China and has quickly become the focus of worldwide attention. Information and discussions about COVID-19 have spread rapidly online, especially on social media.

To fight against COVID-19 and promote the prevention and control of the epidemic, researchers have recently made efforts in various aspects; their research involves epidemiological research ^5,6,7^, diagnostic methods of the novel coronavirus ^8,9,10,11^, clinical characteristics of the disease ^12,13,14,15,16^, characteristics of disease transmission ^17,18,19^, development of candidate therapies ^20,21,22^, the identification of animal hosts ^23,24,25,26^, etc. However, there has been no research conducted on the public’s attention to COVID-19. Since public participation is required to prevent and control the epidemic spread of infectious diseases, it is extremely important to learn about the public’s attention to COVID-19 during the current epidemic. Such knowledge is of great significance when guiding people to respond appropriately to the epidemic and helping them learn how to cope with the sudden infectious disease of COVID-19, and it also supports social stability ^27^.

Social media has developed rapidly in recent years. Increasing numbers of public health departments and individuals are using social media platforms to communicate and share information during public health emergencies. Social media has become an important channel for promoting risk communication during the crisis^28,29^. The use of social media to measure public attention has also been gradually applied to research on infectious diseases, such as H7N9^27,30,31^, Ebola^28,32,33, 34,35,36^, Zika virus^29, 37,38^, MERS-CoV^39^, and Dengue fever^40^.

Sina Microblog (“Weibo” for short) is one of the most popular social media platforms in China and is equivalent to Twitter within China. As of the fourth quarter of 2018, the number of monthly active users had reached 462 million, and approximately 200 million people are using Sina Microblog every day^41^. The Sina Microblog hot search list is the ranking of the most followed and hottest information on Sina Microblog and is the most popular functional module in Sina Microblog applications^42, 43^. This ranking is sorted according to the search volume of the hot topics that users searched for within a certain period. The higher the search volume is, the higher the ranking is, which directly reflects the public attention and attitude toward the topic.

This paper studies public attention given to COVID-19 on Sina Microblog by searching for and analyzing topics related to the COVID-19 epidemic on the Sina Microblog hot search list from December 31, 2019, to February 20, 2020. This is the first time that a Chinese social media platform (i.e., Sina Microblog) has been used to study the public attention of COVID-19^44^. In this study, we describe the trend of public attention given to topics related to the COVID-19 epidemic and the hot topic keywords of public concern, we analyze the emotional tendencies and trends of hot topics related to the COVID-19 epidemic, and we conduct a visual cluster analysis of the hot topic content. This approach is used to obtain timely access to public responses so that the government and the health department can better communicate with the public on health issues and take appropriate measures to prevent and control the epidemic ^35, 39^.

## Methods

### Research Overview

The research process mainly includes five steps: (1) collect topics related to the COVID-19 epidemic on the Sina Microblog hot search list, (2) segment the collected text into words, (3) describe the Sina Microblog search trend about the COVID-19 epidemic; (4) evaluate public opinion through word frequency and sentiment analysis; and (5) construct a social network of public opinion through the subject analysis of public opinion content. Each step is described in detail below.

### Data Collection

We obtained information on the COVID-19 infection in mainland China from the National Health Committee of the People’s Republic of China (PRC NHC) ^45^. The Wuhan Municipal Health Committee first reported viral pneumonia of unknown causes on December 31, 2019 ^46^. This study collected topics on the Sina Microblog hot search list from December 31, 2019, to February 20, 2020, using Weibo Search Rank^47^ from ENLIGHTENT^48^, and selected topics and search volume related to the COVID-19 epidemic. A total of 4,056 topics related to the COVID-19 epidemic were on the hot search list, and 3,234 remained after excluding duplicate topics; this group was used as the data basis for further processing, analysis, and discussion.

### Data Processing

The topics related to the COVID-19 epidemic on the Sina Microblog hot search list from December 31, 2019, to February 20, 2020, were summarized and classified daily in chronological order. We used the Chinese words segmentation function in ROST CM6.0 (ROST Content Mining System Version 6.0) to segment the content of hot-search topics on Sina Microblog into words ^49^. After word segmentation, the text was processed by merging synonyms and deleting nonsense words to provide a basis for subsequent research.

### Data Analysis

### Trend Analysis

We plotted the number of topics related to the COVID-19 epidemic on the Sina Microblog hot search list and the cumulative search volume by date to explore the public’s attention given to the COVID-19 epidemic over time. Pearson correlation analysis was used to explore the relationship between the number of hot search topics related to the COVID-19 epidemic and the cumulative search volume.

### Word frequency and sentiment analysis

After Chinese word segmentation and invalid word filtering, we used ROST CM6.0 software to perform word frequency statistics and sentiment analysis on the Sina Microblog hot topics related to the COVID-19 epidemic. We calculated the frequency of keywords appearing in the hot search topics on Sina Microblog, explored the sentiment tendency of each hot search topic to the COVID-19 epidemic, and calculated the emotional score.

### Social network analysis and visualization

The high-frequency keywords and their frequencies were extracted from the Sina Microblog hot topic text after word segmentation and invalid word filtering. We used VOSviewer software to create a visual knowledge map of keyword co-occurrence analysis and cluster analysis, using the keyword co-occurrence frequency as the weight^50^.

## Results

### Search trend of COVID-19 epidemic on Sina Microblog hot search list

Figure 1 lists the number of COVID-19 epidemic-related topics and the cumulative search volume on the Sina Microblog hot search list from December 31, 2019, to February 20, 2020. Pearson correlation analysis shows that the number of topics related to the COVID-19 epidemic is positively correlated with the cumulative search volume of the topic per day (Pearson correlation coefficient = 0.767, P= .000). In other words, the more topics are listed, the greater the cumulative search volume related to the COVID-19 epidemic per day. We can see that the public’s attention to the COVID-19 epidemic on Sina Microblog can be divided into three stages. Stage A (December 31, 2019, to January 18, 2020) has low and unstable public attention, which represents the incubation period. Stage B (January 19 to 26, 2020) has a concentrated increase in public attention, which represents the epidemic period. Stage C (January 27 to February 20, 2020) demonstrates continued public attention to the epidemic, representing a widespread period.

**Figure 1.**
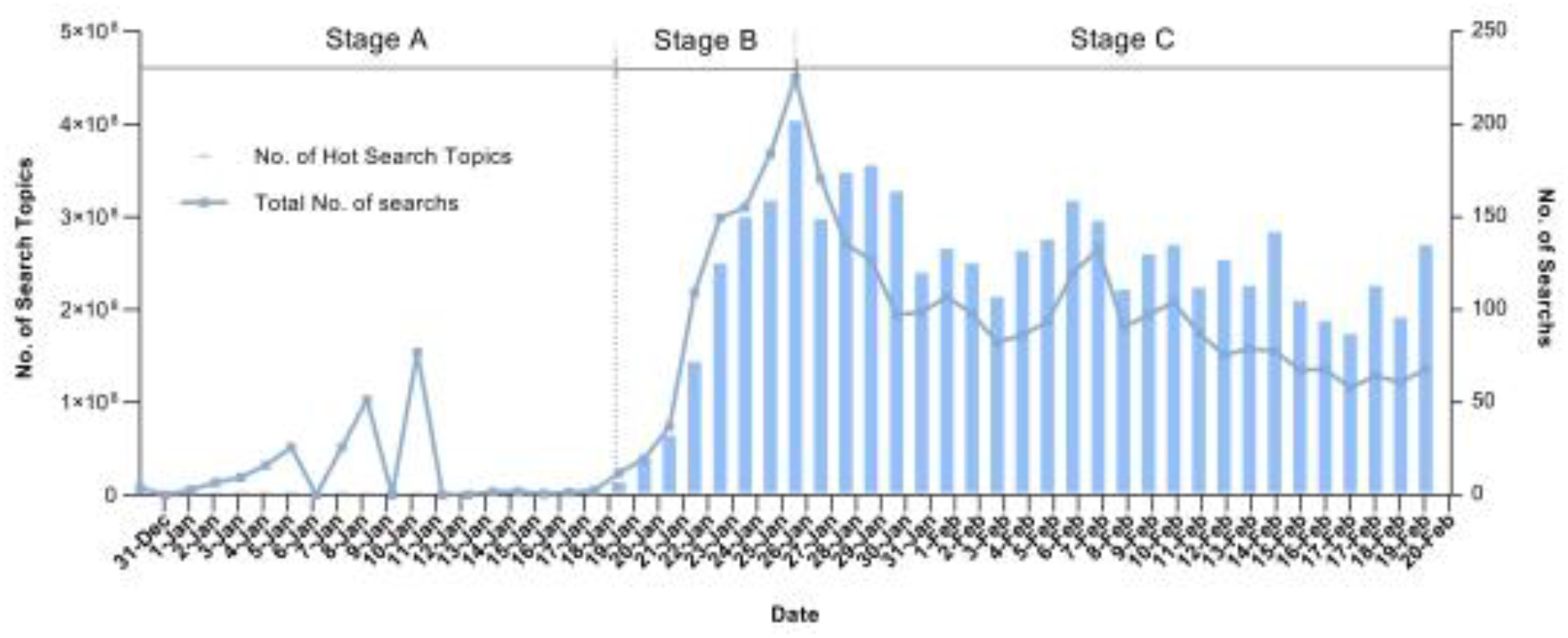
From December 31, 2019, to February 2, 2020, the number of topics and cumulative search volume of the COVID-19 epidemic on the Sina Microblog hot search list.

### Frequency and distribution of hot search keywords related to the COVID-19 epidemic on Sina Microblog

The top 15 keywords and their frequencies for the three stages of the public’s attention to the COVID-19 epidemic are shown in Table 3. We can find that “Wuhan”, “case” and “pneumonia” always appear in three periods as hot keywords, and the remaining keywords in the different periods are slightly different. In stage A, “unknown cause” and “new coronavirus” are the main keywords, indicating that in the initial stage of the epidemic, viral pneumonia had just been discovered, and the cause was unknown. After the pathogen was initially identified as a new coronavirus, the public began to search for information on new coronaviruses to learn relevant knowledge. Compared with stage A, stage B has new keywords such as “new”, “mask”, “first case”, and “first-level response”. The reason for this is that the COVID-19 epidemic had spread across the country during this stage. The first cases had appeared successively throughout the country, and the number of confirmed cases was increasing. The outbreak of the epidemic made the people and the government aware of the importance of prevention. People began to buy masks, and governments at all levels initiated first-level responses to major public health emergencies. Compared with the previous two stages, the main keywords appearing in stage C are “discharged”, “national”, “materials” and “Huoshenshan Hospital”. At this stage, the public’s attention had shifted to material donation and medical service assistance in the key epidemic areas in Wuhan. Moreover, the epidemic had spread throughout the country, and the public was more concerned about the rehabilitation of patients.

### Sentiment analysis of the hot search topics related to the COVID-19 epidemic on the Sina Microblog

Afer segmentation, we imported the Sina Microblog hot topic text related to the COVID-19 epidemic into the ROST6.0 sentiment analysis tool and obtained the sentiment and proportion of the three stages of public attention given to the topics related to the COVID-19 epidemic on Sina Microblog (Figure 2). Emotions are classified as positive, negative, and neutral emotions. Based on this, positive and negative emotions are subdivided into three categories: general, moderate, and high. Neutral emotions are not subdivided. We found that the sentiment of the hot topics of the COVID-19 epidemic on Sina Microblog in stage A tended to be negative, accounting for 29.17%, of which high and moderate negative emotions accounted for 4.17% and 16.67%, respectively, and positive emotions accounted for the lowest proportion at 8.33%. In stage B, the majority of the hot search topics were neutral; however, 22.85% of the hot search topics showed negative emotions, with 0.31% showing high negative and 3.60% showing moderate negative, while positive emotions accounted for 12.52%. In stage C, 15.44% of the hot search topics showed positive emotions, of which moderate positive emotions accounted for 2.26% and highly positive emotions accounted for 0.31%; in addition, 24.33% of the hot search topics showed negative emotions. Based on the comprehensive analysis of the three-stage emotional tendencies, the public’s negative emotions toward the COVID-19 epidemic were weakened as a whole, and their positive emotions were generally enhanced.

**Figure 2.**
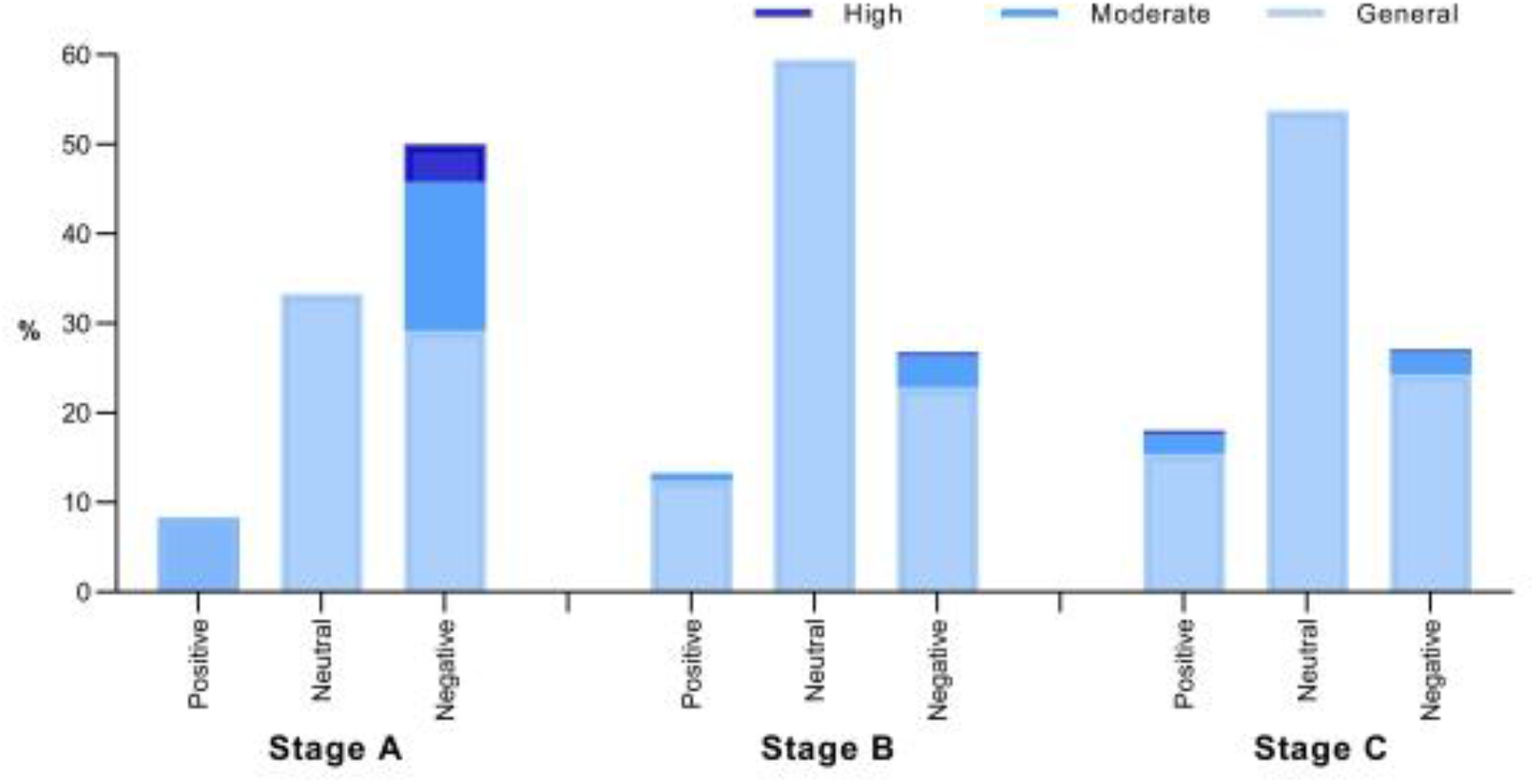
The sentiment statistics of the Sina Microblog hot search topics related to the COVID-19 epidemic.

Figure 3 shows the trend of the proportion of the daily emotional tendencies of the hot search topics related to the COVID-19 epidemic from December 31, 2019, to February 20, 2020. We can see that the three kinds of emotions are relatively unstable before January 9, 2020. From January 9 to January 20, 2020, negative emotions accounted for the largest proportion, followed by neutral emotions, and positive emotions accounted for the smallest proportion.

**Figure 3.**
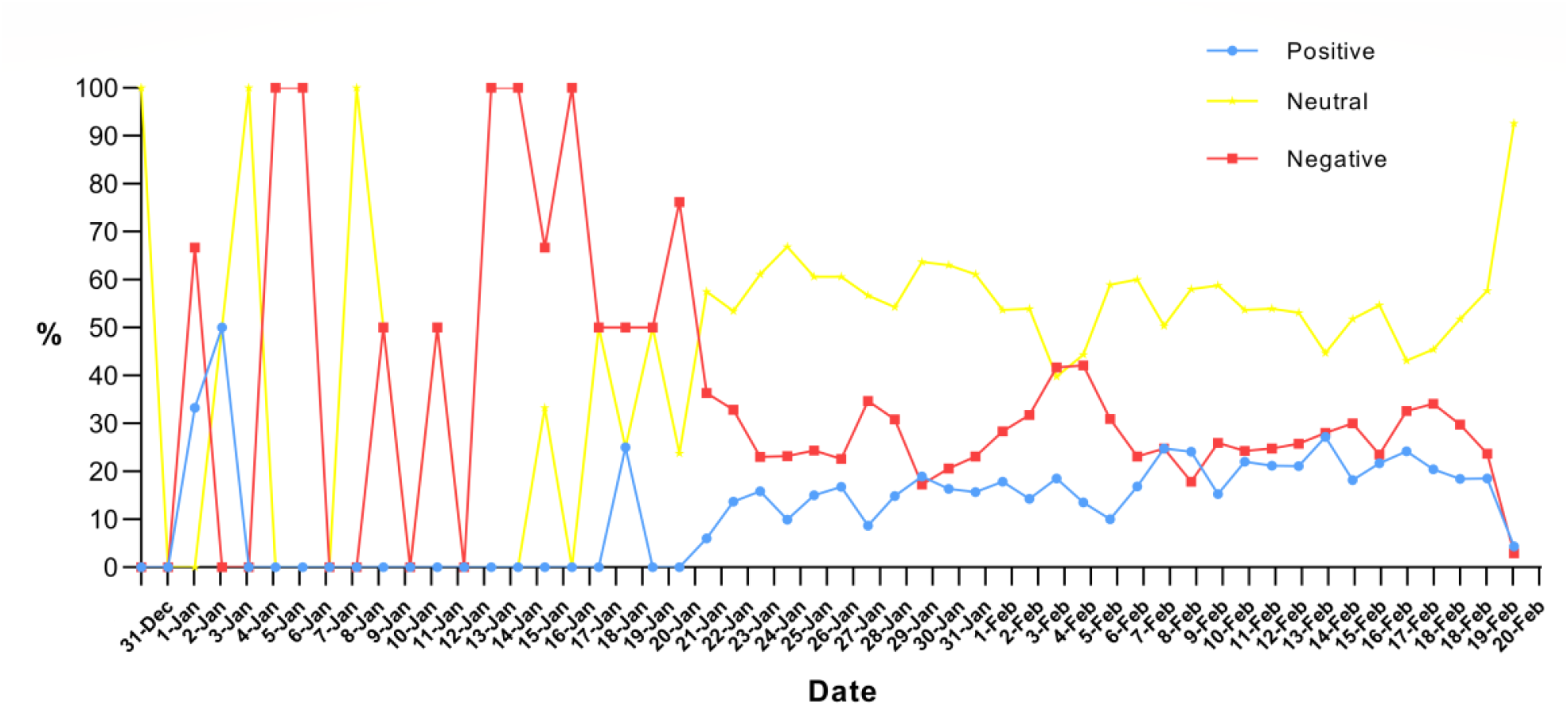
The sentiment trend of hot search topics related to the COVID-19 epidemic from December 31, 2019, to February 2, 2020.

Compared with before January 20, 2020, the positive emotions of the hot search topics related to the COVID-19 epidemic after January 20, 2020 are generally on the rise, the negative emotions decline on the whole, and the emotions tend to be stable, as seen in Figure 3. This outcome shows that as the COVID-19 epidemic began to spread throughout the country after January 20, 2020, the public eased their concerns and fears caused by their uncertainty toward and ignorance of the epidemic and responded to the COVID-19 epidemic with a more objective attitude.

### Social semantic network analysis of hot search topics related to the COVID-19 epidemic on Sina Microblog

To explore the themes reflected by the related topics of the COVID-19 epidemic on the Sina Microblog hot search list, this study used VOSviewer to generate clusters and co-occurrence networks of topic keywords. The results are shown in Figure 4. In the figure, the larger the size of the nodes and the font, the greater the weight of the keyword is, and the more it is in the core position. The connection between the nodes indicates that the keywords on the two nodes have appeared together. The thicker the connection is, the higher the frequency of co-occurrence and the closer the connection between the nodes. Nodes with the same color belong to the same cluster.

**Figure 4.**
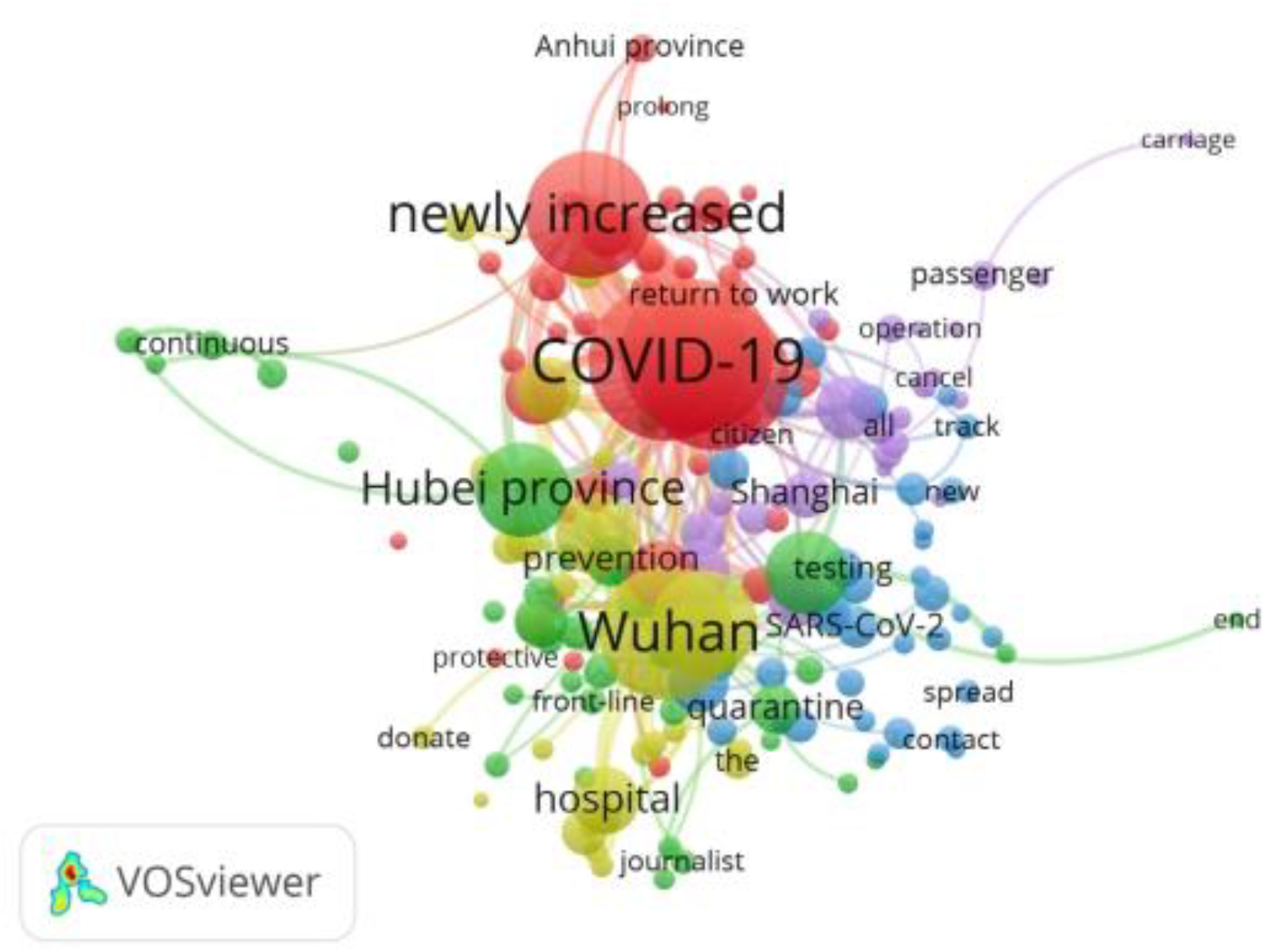
Social network of high-frequency keywords in the Sina Microblog hot search topics related to the COVID-19 epidemic.

**Table 1.** Top 15 keywords about the COVID-19 epidemic on the Sina Microblog hot search list in the three periods.

| **Stage A** |  | **Stage B** |  | **Stage C** |  |
| --- | --- | --- | --- | --- | --- |
| Dec 31, 2019, to Jan 18, 2020 |  | Jan 19 to Jan 26, 2020 |  | Jan 27 to Feb 20, 2020 |  |
| **Key Word** | **Freq** | **Key Word** | **Freq** | **Key Word** | **Freq** |
| Wuhan | 17 | COVID-19 | 141 | Wuhan | 316 |
| pneumonia | 14 | Wuhan | 124 | COVID-19 | 283 |
| unknown cause | 9 | confirmed diagnosis | 87 | case | 212 |
| novel coronavirus | 6 | case | 84 | confirmed diagnosis | 212 |
| patient | 5 | add | 52 | add | 199 |
| case | 5 | masks | 37 | epidemic | 190 |
| Thailand | 4 | epidemic | 31 | Hubei | 159 |
| leave hospital | 2 | Hubei | 29 | masks | 149 |
| death | 2 | startup | 25 | patient | 149 |
| person-to-person spread | 2 | first case | 22 | hospital | 98 |
| add | 2 | pneumonia | 20 | leave hospital | 90 |
| Japan | 2 | Beijing | 20 | nationwide | 83 |
| COVID-19 | 2 | patient | 19 | Beijing | 20 |
| epidemic disease | 2 | first-level response | 18 | goods and materials | 59 |
| eliminate | 2 | novel coronavirus | 17 | Huoshenshan hospital | 59 |

<sup>a</sup> Abbreviations: Freq, Frequency; COVID-19, coronavirus disease - 2019.

According to the network visualization graph constructed in Figure 4, we can see that the keyword “COVID-19” is at the core node position, and the two nodes “Wuhan” and “add” are next. The core topic of public concern about the COVID-19 epidemic is COVID-19 itself, and the public was extremely concerned about the status of the epidemic in Wuhan and the new cases. We can divide the high-frequency keywords of topics into the following five clusters:

Cluster 1 (Red Cluster): Discussion on the new cases, the outbreak of COVID-19 across the country and the impact of the epidemic on the resumption of school and work. The keywords included are “case”, “newly increased “, “appear”, “Hubei Province”, “Beijing”, etc. (Figure 5a).

Cluster 2 (Green Cluster): Search for news reports on the frontline of the epidemic and related measures for prevention and control. The keywords include “Wuhan”, “front-line”, “inpatient”, “quarantine”, “living at home”, etc. (Figure 5b).

Cluster 3 (Blue Cluster): Search for interpretations of the epidemic situation and prevention and control knowledge of experts and relevant health departments, as well as the discussion on the source of infection. The keywords in this cluster include “academician”, “WHO”, “face mask”, “epidemic prevention”, “symptom”, “virus”, and “novel coronavirus”. (Figure 5c).

Cluster 4 (Yellow Cluster): Search for frontline medical services such as frontline hospital construction and medical team support. This cluster includes the keywords “patient”, “hospital”, “Leishenshan”, “Huoshenshan”, “first batch”, etc. (Figure 5d).

Cluster 5 (Purple Cluster): Search for the global spread of the disease and searching online for fellow passengers with confirmed cases. This cluster contains the keywords “passenger”,“carriage”,“cruise”,“suspend”, “infection”, “Japan”, etc. (Figure 5e).

**Figure 5a.**
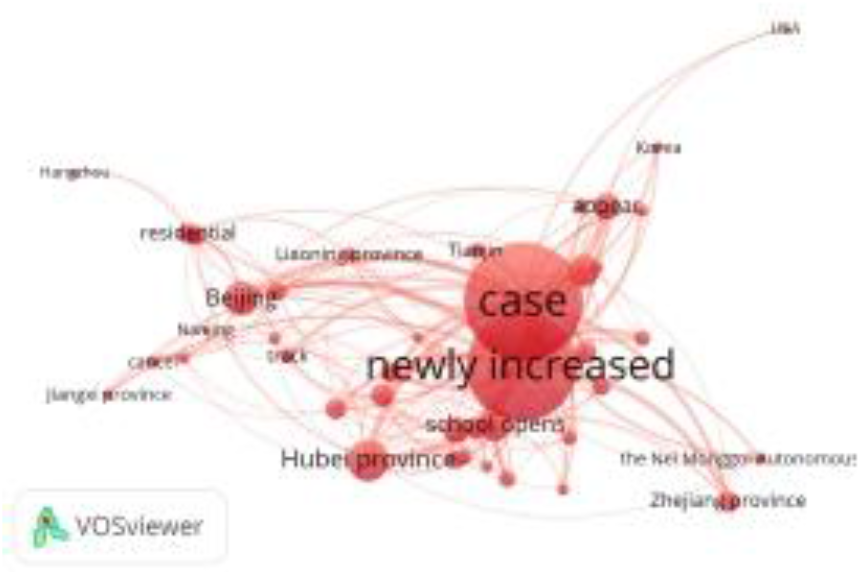
Cluster 1

**Figure 5b.**
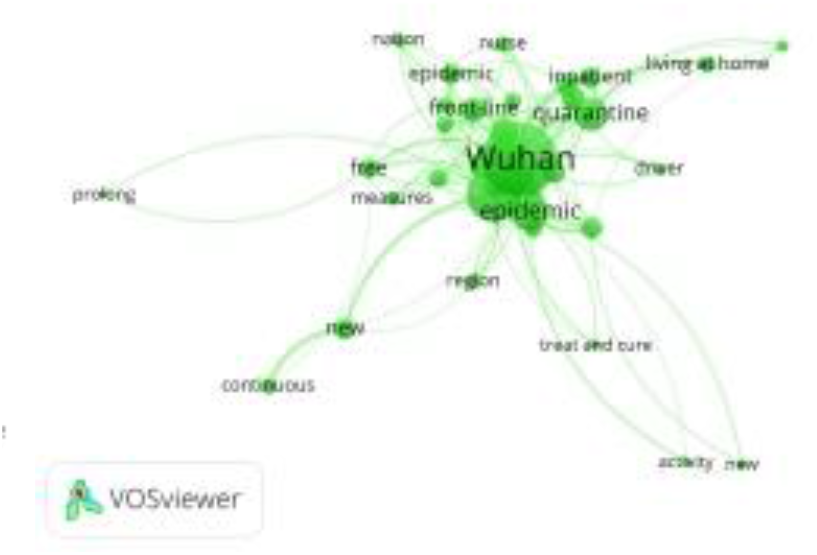
Cluster 2

**Figure 5c.**
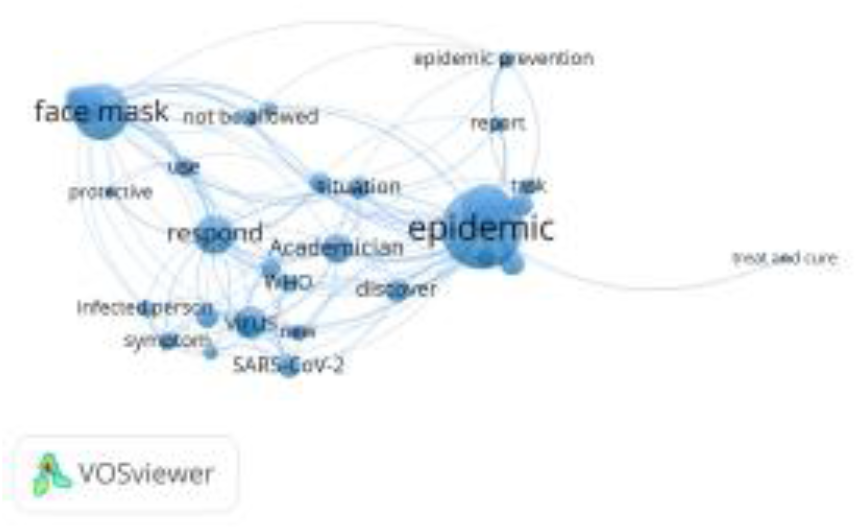
Cluster 3

**Figure 5d.**
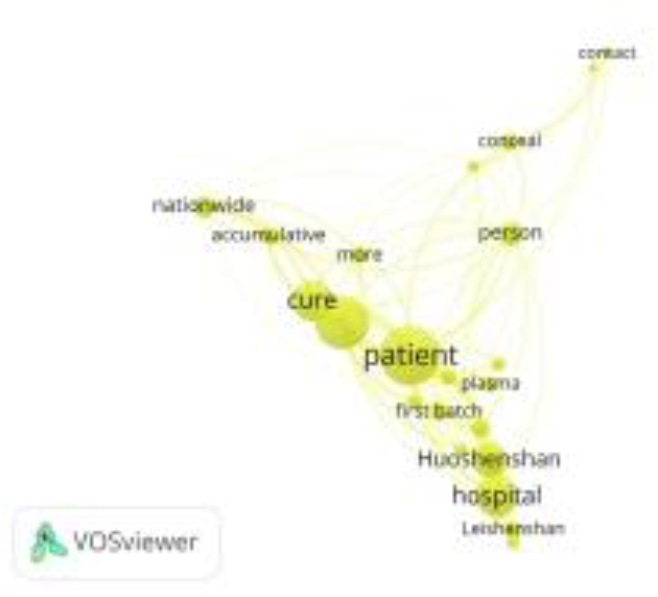
Cluster 4

**Figure 5e.**
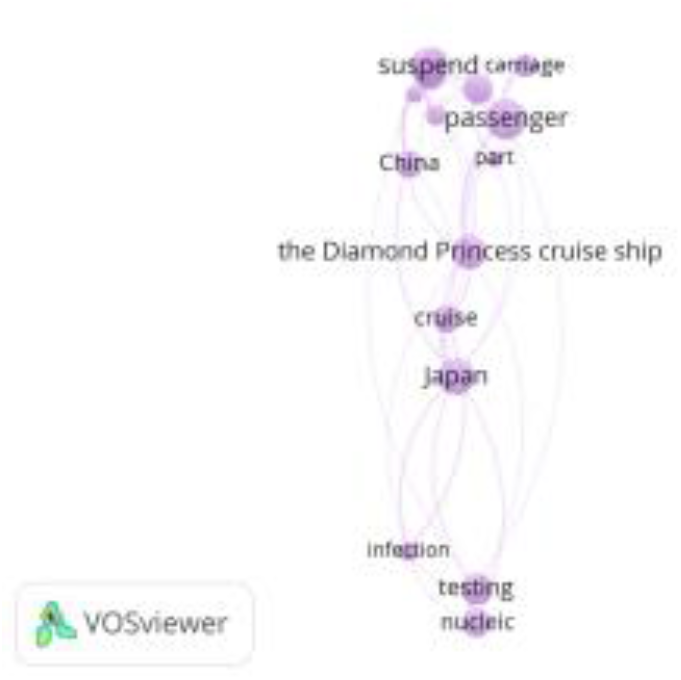
Cluster 5

## Discussion

### Principal Results

In recent years, as people increasingly seek health information on social media, social media has played an increasingly important role in public health emergencies^29,35^. However, there have been no relevant studies that have incorporated social media analysis into the public response to the COVID-19 epidemic. We used the Sina Microblog hot search list to analyze the public’s attention to the COVID-19 epidemic from December 31, 2019, to February 20, 2020, which was 52 days after the first disclosure of COVID-19 by the Chinese health department. There are four main findings from this study. First, we analyzed the changing trend of public attention given to the COVID-19 epidemic, which can be divided into three stages. Second, the hot topic keywords of the public’s attention at each stage are slightly different. In addition, the public’s emotional tendency toward the hot search topics related to the COVID-19 epidemic changed from negative to neutral across the study period. As a whole, negative emotions weakened, and positive emotions increased. Finally, we divided the topics of public concern about the COVID-19 epidemic into five categories through social semantic network analysis. This study analyzes the public’s response to and attention given to the COVID-19 epidemic, which will help public health professionals monitor public response, identify public needs as early as possible, make timely public health prevention and control measures, and disseminate knowledge to citizens in a targeted manner to better respond to the current COVID-19 epidemic.

From December 31, 2019, to February 20, 2020, the public’s attention given to the COVID-19 epidemic on Sina Microblog can be divided into three stages. In the beginning, there was little public attention paid to the epidemic, and then the concentration of attention increased. Next, the public’s attention given to the epidemic generally declined, but continued attention was still paid. In the first stage (December 31, 2019, to January 18, 2020), the Chinese public paid less attention, as pneumonia was only reported in Wuhan, China. On January 19, 2020, suspected cases appeared in Shanghai and Shenzhen, and body temperatures began to be measured at the Wuhan Airport and Railway Station. People began to notice the severity of the epidemic, and the level of attention paid to the COVID-19 epidemic began to increase until January 26, 2020. We can see that when the epidemic began to spread across the country, the public responded quickly to news of the COVID-19 outbreak on Sina Microblog^39^. This is because COVID-19 is a new infectious disease, which means that no effective treatment has been found and no corresponding vaccine has been developed. When the outbreak began, the public was eager to search the relevant knowledge and information online to meet their own protection needs^27,30^. In the third stage (January 27 to February 20, 2020), although the number of cases of COVID-19 was still increasing, the number of Sina Microblog topics and search volume was decreasing. The reason for this may be because the epidemic information was relatively saturated, and it was difficult for people to acquire more new knowledge through a Sina Microblog search^27^. It may also be that people are no longer paying close attention to the epidemic dynamic to gain a sense of security as time goes on, and the public’s consciousness tends to be rational^51^.

The keywords for the three different stages of the public’s attention to the COVID-19 epidemic are slightly different. From the hot search keywords of each stage, we found that the public did not know much about the virus and its causes in the first stage. The main keywords of the search to seek relevant knowledge were “unknown cause” and “novel coronavirus”. During the second stage, the epidemic began to spread throughout the country, the first cases were reported in different areas successively, and the number of confirmed cases continued to increase, thereby making the public and government aware of the importance of prevention. During the third stage, the epidemic spread widely throughout the country; Wuhan was the most seriously affected area, and the city was locked down. The public’s attention was mainly shifted to material donation and medical assistance in Wuhan.

Compared with a qualitative research, it is more accurate to obtain the hot spots of public attention through keyword frequency analysis, as this process has higher accuracy and credibility in the research of hot spots and their development trends^40^. Based on our high-frequency keyword analysis of the Sina Microblog topics related to the COVID-19 epidemic, we can obtain information about the concerns and opinions of Sina Microblog users at different stages^52^. Studies have found that people’s interest in infectious diseases on social media is linked to the latest news and major events. Studies have also shown that people will pay attention to and search for disease-related words as the spread of infectious diseases changes ^30^.

According to the sentiment tendencies of the hot search topics on the COVID-19 epidemic on Sina Microblog, the first stage of emotion was negative, and the second and third stages were neutral. On the whole, negative emotions weakened, and positive emotions increased. Previous studies have pointed out that there is also an important relationship between emotions and content on social media^53^. The content analysis of social networks can identify people’s attitudes or reactions to specific health hazard events^30,39^. In the first stage, there was less public attention given to the Sina Microblog hot search list. Most of the topics related to the COVID-19 epidemic were about the notification of pneumonia and the virus, and the emotions tended to be negative. At that time, the public had a strong demand for information on public health emergencies such as infectious diseases. When the information demand could not be fully satisfied, the users’ emotions were negative^40^. As the epidemic progressed to the second and third stages, the public sentiment tended to be neutral because increasingly more news was reported at this stage, and objective events became the mainstream information on the Sina Microblog hot search list. The public reduced their previous levels of worries and fears about the epidemic; their negative feelings weakened, and their positive emotions increased.

More hot search topics mentioned information about prevention or protection, which is conducive to public health communication and promotion.

We divided the COVID-19 topics with the highest levels of public concern into five categories: (1) new COVID-19 epidemics and their impact, (2) frontline reporting of the epidemic and prevention and control measures, (3) expert interpretation and discussion on the source of infection, (4) medical services on the frontline of the epidemic, and (5) focus on the global epidemic and look for suspected cases. From the search of the subject content, we can see that during the outbreak of the novel coronavirus, the public, the news media, and the health department all actively used Sina Microblog as a platform for disseminating COVID-19-related information^29^, indicating that Sina Microblog is a communication channel for both individuals or organizations to publicize COVID-19 symptoms, preventive measures and related policies^37^. In addition, as a real-time and extensive online platform, Sina Microblog provides a channel for information dissemination. For example, in this emergency outbreak, the public has made good use of the platform to find fellow passengers with confirmed cases, thereby playing an important role in preventing and controlling the disease transmission.

## Limitations

There are some limitations to this study. First, our study was limited to the period 52 days after COVID-19 was first disclosed by the Chinese health department; thus, the situation after February 20, 2020, was not included in this study. Second, the data source for this study is relatively narrow. We regard Sina Microblog as the only social media platform in China, excluding other popular social media data sources, such as WeChat and ByteDance, and the study is limited to the publicly available data on the Sina Microblog hot search list. In addition, due to the lack of detailed information about the users who contributed to the search volume of the Sina Microblog hot search list, we could not describe the social demographic information of the Sina Microblog users, and we failed to obtain the geospatial distribution of Sina Microblog active users; thus, we could not calculate the average attention of the public in different regions of China.

## Conclusions

Our study found that social media platforms (i.e., Sina Microblog) can be used to measure public attention given to public health emergencies. Our study shows that a large amount of information about the COVID-19 epidemic was disseminated and received widespread public attention on Sina Microblog during the novel coronavirus epidemic. We have learned about the hotspots of public concern regarding the COVID-19 epidemic on Sina Microblog. These findings can help the government and health departments better communicate with the public about public health and then translate public health needs into practice to create targeted measures to prevent and control the spread of COVID-19.

## Data Availability

Data used and analyzed in the study are available from public resources.

## Acknowledgments

Not applicable.

## Abbreviations

COVID-19: Corona Virus Disease 2019

